# Between Geography and Demography: Key Interdependencies and Exit Mechanisms for Covid-19

**DOI:** 10.1101/2020.04.09.20059592

**Authors:** Antonio Scala, Andrea Flori, Alessandro Spelta, Emanuele Brugnoli, Matteo Cinelli, Walter Quattrociocchi, Fabio Pammolli

## Abstract

We develop a minimal compartmental model to analyze policies on mobility restriction in Italy during the Covid-19 outbreak. Our findings show that a premature lockdown barely shifts the epidemic in time: moreover, beyond a critical value of the lockdown strength, an epidemic that seems to be quelled fully recovers after lifting the restrictions. We investigate the effects on lockdown scenarios and exit strategies by introducing heterogeneities in the model. In particular, we consider Italian regions as separate administrative entities in which social interactions through different age classes occur. We find that, due to the sparsity of the mobility matrix, epidemics develop independently in different regions once the outbreak starts. Moreover, after the epidemics ha started, the influence of contacts with other regions becomes soon irrelevant. Sparsity might be responsible for the observed delays among different regions. Analogous arguments apply to the world/countries scenario. We also find that disregarding the structure of social contacts could lead to severe underestimation of the post-lockdown effects. Nevertheless, age class based strategies can help to mitigate rebound effects with milder strategies. Finally, we point out that these results can be generalized beyond this particular model by providing a description of the effects of key parameters on non-medical epidemic mitigation strategies.

## 1 Introduction

Epidemic modelling is not a new field, and, especially in the veterinary field, scientists have developed models for diseases and infections that have been successfully applied to mitigate their impact. However, when coming to humans, epidemic modelling has never had such a great impact as we are experiencing now with Covid-19.

Epidemic models can vary in complexity; more complex models require highquality granular data. However, even if each single model may risk to oversimplify the underlying system and may lack crucial aspects, different modelling approaches can provide different insights and contribute to the decision process for the policies to be applied [1]. Despite the sophistication of epidemic models, they often still lack important aspects, like the failure to include economic aspects (see [2, 3, 4]) or, in the case of human epidemics, to assess the influence of media communication [5]. At the moment, although the World Health Organization (WHO) organizes regular calls for Covid-19 modelers to compare strategies and outcomes, policymakers find difficult to handle the discrepancies between the proposed models^1^: while uncertainty and complexity are accustomed traveling companions for scientists, they are possibly the worst scenario for decision makers.

For containing the Covid-19 epidemic, governments worldwide have adopted severe social distancing policies, ranging from partial to total lockdown of their population [6]. These restrictions have led to the disruption of business activities in many economic sectors and areas. Furthermore, the age class of 15–64 years, those engaged in the labor force, account for the majority of Covid-19 infections [7]. Hence, the impact of contagion and lockdown measures on economic activities appear to be substantial and pervasive. As a consequence, disposable incomes will drop significantly during the outbreak; consumption by households will also decrease and the closure of schools could result in lost education with long-term effects on human capital accumulation. Thus, the socio-economic implications of the Covid-19 are likely to play a detrimental impact not only on public health systems, but also on several production sectors and retail markets and, possibly, on wider global value chains.

Starting from such premises, we apply a model-based scenario analysis for Covid-19, highlighting the role non-medical variables play in the epidemic spreading and how those dimensions are likely to impact on lockdown policies and exit (from lockdown) strategies [8]. In particular, we study how to schedule shortterm interventions in response to an emerging epidemic when geographic and demographic variables are accounted in the model. We are indeed interested in analyzing how mobility restriction measures and the timing of the lockdown release might affect the total fraction of infected, the peak prevalence, and, possibly, the delay of the epidemic when model heterogeneity is driven by regional boundaries and age classes [8]. To such an end, we use a very general compartmental model, minimally adapted to describe Italian data, and we analyze the effect of disregarding regional compartmentalization and age classes. Notice that our results are general for the vast class of epidemic models where transmission rate is proportional to the number of susceptible people times the density of infected. In a nutshell, our scope is to understand which dimensions, beside medical ones, should be considered and available to leverage strategies for containing epidemic even before having detailed quantitative predictions. Secondly, we also aim to point out how disregarding such dimensions in epidemic models could seriously affect exit scenarios.

Our approach contributes to the extant literature on the trade-offs between a mitigation scenario, aimed to slow down the epidemic contagion, and a suppression scenario, consisting on lowering the risk of contagious while maintaining it indefinitely [8, 9, 10]. Our projections show that an anticipated lockdown just shifts the epidemic in time, with a delay proportional to the anticipation time and grows with the strength of the lockdown. Moreover, beyond a critical lockdown strength, while at the beginning the epidemic seems to be quelled, it fully recovers its strength as soon as the lockdown is lifted. If we consider regional mobility flows, due to the sparsity of the mobility matrix, epidemics develop independently in different regions once the epidemic is started. We postulate that the sparsity of the inter-regional mobility matrix could be responsible of the delays observed among different regions and that the same consideration may apply to explain the independent dynamic of the epidemic spreading in different countries. Finally, if we add social contact heterogeneity on different age classes we find that the structure of social contacts is of primarily importance for avoiding an underestimation of the post-lockdown effects and that age class based strategies can help to mitigate rebound effects with milder strategies.

## 2 Model

To analyze mobility-restrictive policies, we set up a minimal compartmental model [10, 11]. Although many models, both mechanistic and stochastic, have been proposed for the Covid-19 infection, it is very difficult to assess their effectiveness since the data collected from the national healthcare systems suffer from the lack of homogeneous procedures in medical testing, sampling and data collection [12]. Even so, there is still the problem of assessing the impact of the variability in social habits during the epidemics [10, 13].

Especially in the early phases of the epidemic – i.e. the ones characterised by an exponential growth – different models sharing the same reproduction number *R*_0_ can fit the data with a similar accuracy (see Sec. 8.1). Since we are interested in the qualitative scenarios and not in the detailed predictions, we adapt the *SIR* model, the most basic epidemic model for flu-like epidemics, to the observed data available in the Italian case.

In our model, we rely on four compartments, namely: *S, I, O, R*. Hence, *S*(usceptible) individuals can become *I*(nfective) when meeting another infective individual, *I*(nfectives) either become *O*(bserved) – i.e. present symptoms acute enough to be detected from the national health-care system – or are *R*(emoved) from the infection cycle by having recovered; also *O*(bserved) individuals are eventually *R*(emoved) from the infection cycle (see Fig. 1 for a visual representation of the model workflow). Notice that, it is not still clear if there is an asymptomatic phase [14, 15]; in our model, we are implicitly assuming that asymptomatics are infective and their recovery time is the same of the *I* class. Hence, our model is described by the following differential equations:

**Fig. 1.**
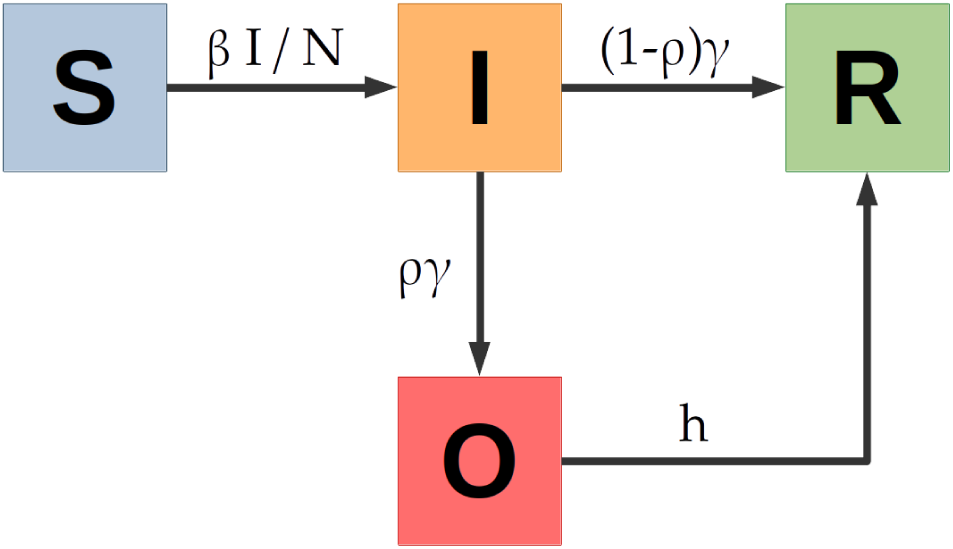
The *SIOR* compartimental model: workflow of the epidemic process. A *S*(usceptible) individual becomes *I*(nfective) when meeting an infectived person. An *I*(nfectived) either become *O*(bserved), with symptoms acute enough to be detected from the national health-care system, or is *R*(emoved) from the infection cycle by having recovered. An *O*(bserved) individual can also be *R*(emoved) from the infection cycle having become immune. The parameter *β* defines the rate at which a susceptible becomes infected, *γ* represents the rate at which infected either become observable or recover, *ρ* is the fraction of infected that become observed from the national health-care system and *h* is the rate at which observed individuals are removed from the infection cycle.

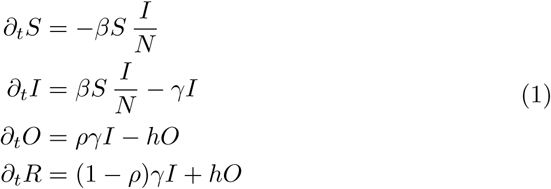

Here, *N* = *S* + *I* + *O* + *R* is the total number of individuals in a population, the transmission coefficient *β* is the rate at which a susceptible becomes infected upon meeting an infected individual, *γ* is the rate at which an infected either becomes observable or is removed from the infection cycle. Like the *SIR* model, the basic reproduction number is *R*_0_ = *β/γ*; the extra parameters of the *SIOR* model are *ρ*, the fraction of infected that become observed from the national health-care system, and *h*, the rate at which observed individuals are removed from the infection cycle. Notice that we consider that *O*(bserved) individuals not infecting others, either because hospitalized or because in a strict quarantine.

## 3 Lockdown

The Italian lockdown measures of the 8^th^ and 9^th^ of March [16, 17] were intended to change the mobility patterns and to reduce the de-visu social contacts, thanks both to quarantine measures and to an increased awareness of the importance of social distancing. The analysis of Facebook mobility data^2^ [18] confirms that the lockdown has reduced both the travelled distance and the flow of travelling people.

Let us consider the effects of such lockdown measures on the parameters of our model. The rate *γ* is the most unaffected, since it is related to the “medical” evolution of the disease; analogous arguments apply to the probability *ρ* of developing a condition serious enough to be observed (although variations in testing schemes and alert thresholds could influence such parameter) and to the rate *h* of exiting such a condition. On the other hand, the transmission coefficient *β* can be thought as the product of a contact rate times a disease-dependent transmission probability. Hence, if we assume that the speed of Covid-19 mutation is irrelevant on our timescales, lockdown strategies mostly influence *β* by reducing the contact rate among individuals.

To adapt the *SIOR*’s parameters to the Italian data [19], we compare the reported cumulative number of Covid-19 cases *Y* ^Obs^ with the analogous quantity *Y* ^model^ = *∫ργIdt* in our model. We first estimate model’s parameters by least square fitting on the pre-lockdown period. Since in such range the data *Y* ^Obs^ show an exponential growth trend, we are possibly observing a very early phase of the epidemic, where *β−γ* equals the growth rate of *Y* ^Obs^ (see Sec. 8.1).

In response to the outbreak of Covid-19, a series of estimates of model parameters have been proposed in the literature. Nevertheless, they still show a certain amount of uncertainty about some fundamental variables of the epidemic contagion. Following extant literature [20, 21, 22, 23], we use an infection time duration *τ*_*I*_ *∼*10 *days* and 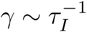. Notice that, while the *R*_0_ would vary linearly with *τ*_*I*_ (i.e. *R*_0_ *∈* [2.5, 4.5] for *τ*_*I*_ *∈* [5, 15], see Sec. 8.1), the scenarios are much less sensitive to such variations. Moreover, we consider an hospitalization period *τ*_*H*_ *∼*10 *days* to estimate 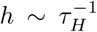. Lastly, we estimate the attenuation *α* of the transmission parameter *β* by refitting the parameters in the post-lockdown time range.

To have an idea of the variability of our parameters [20, 22, 23], we both estimate the variability through a bootstrap procedure and through the sensibility to the variations of *ρ*. While the variations of *ρ* is the factor that mostly shifts the parameter, for our policy-modelling scopes we find a good stability of the fitted parameters even varying *ρ* in a range [10% … 100%]. Namely, we find *β* = 0.35*±*0.01 *day*^*−*1^, *γ* = *h* = 10^*−*1^ *day*^*−*1^, *R*_0_ = 3.5*±*0.1 *day*^*−*1^, *α* = 0.49*±*0.01 for *t*_Lock_ = 15 and *t*_0_ = *−*30 *±* 5 days. Being *t*_Lock_ the time of the lockdown implementation and *t*_0_ the starting time of the epidemic (*i*_0_ = *i*(*t*_0_) = 1). In our analysis we set *ρ* = 40%, *R*_0_ = 3.5 *day*^*−*1^ and *α* = 0.49. Notice that the attenuation *α* in the transmission parameter measures the reduction of the contact rate after the lockdown.

## 4 Italian scenarios

To understand the effects of model’s parameter, but also of introducing several heterogeneities in the modelling, we consider several scenarios. First, we analyze the effects of lockdown starting time and lockdown intensity on the post-exit dynamics in the simple case of a *SIOR* model fitted on Italian data. Then, we study the effect of considering Italy as a collection of separate administrative entities (Italian regions) independently evolving; last, we consider the effects of social interactions through different age classes. We want to point out since the beginning that mobility flows [18] and inter-age social mixing [24] lie at the two opposite range of modelling. In fact, the regional social contact matrix is dense (Fig. 2, left panel), indicating that age classes dynamics are strongly coupled. On the other hand, the inter-regional mobility matrix is very sparse (Fig. 2, right panel), indicating that regions have essentially independent dynamics. This can be seen as an example of the decomposability conditions under which partial dynamics may influence the configuration of the system as whole through the relationships connecting different partitions to the aggregate system (see, e.g., [25, 26, 27, 28]).

**Fig. 2.**
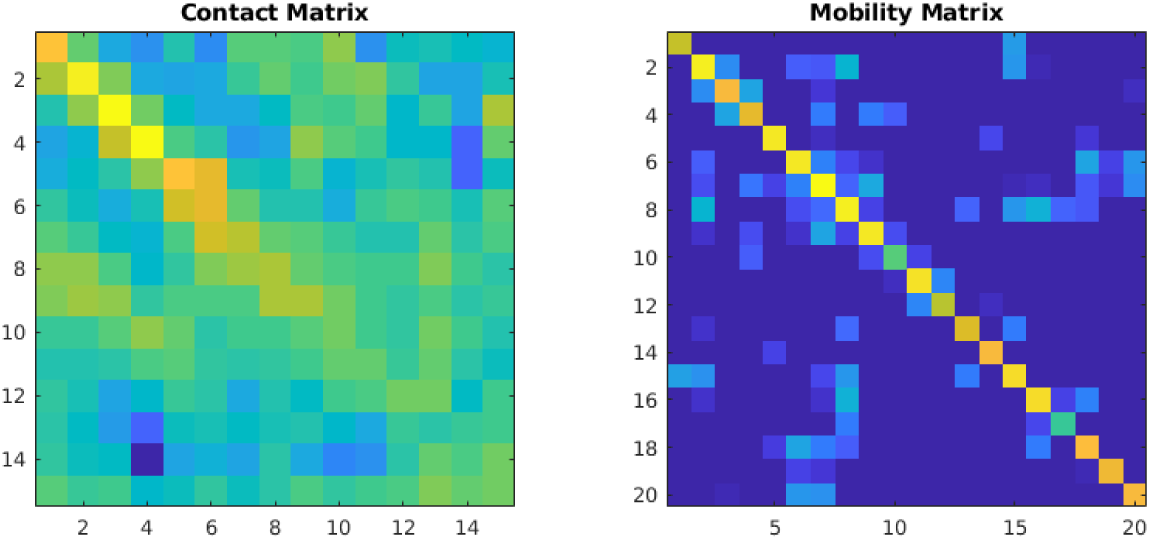
Left Panel: social contact matrix. Right panel: inter-regional mobility matrix. While the regional inter-age social mixing matrix is dense and age classes dynamics are strongly coupled, the inter-regional mobility flows is very sparse; most of the people travel within region.

### 4.1 Simple Exit Strategy

In our model, we first consider a simple exit strategy consisting in releasing the lockdown at a time *t*_Unlock_ after the peak of *O* has occurred. For instance, we imagine that infection proceeds uncontrolled up to time *t*_Lock_; in the following lockdown period [*t*_Lock_, *t*_Unlock_], the transmission coefficient *β* is reduced by a factor *α*; finally, *β* returns to its initial value and herd immunity is responsible for the dampening of the epidemics.

Our results show that the lockdown lowers the peak of *O* - i.e. the individuals with noticeable symptoms - to *∼*70% of the free epidemic one, but also doubles its occurrence time from *∼*1.9 months to *∼*3.8 months, thus an extremely obnoxious effect for the sustainability conditions of the economy of a country. Moreover, since the number of hospitalized patients is a fraction of *O*, much less stress is put on the healthcare system. We can decide to lift the lockdown after the observed people *O* have dropped to a suitable percentage of the maximum peak; as an example, after *∼*4.7 months the peak has reduced to 70% of its initial value, while after *∼*5.2 months to 50%, i.e. *∼*0.5 months later. Notice that, the earlier the lockdown is lifted, the faster *O* decays to zero even if it starts from higher. All such effects are shown in Fig. 3.

**Fig. 3.**
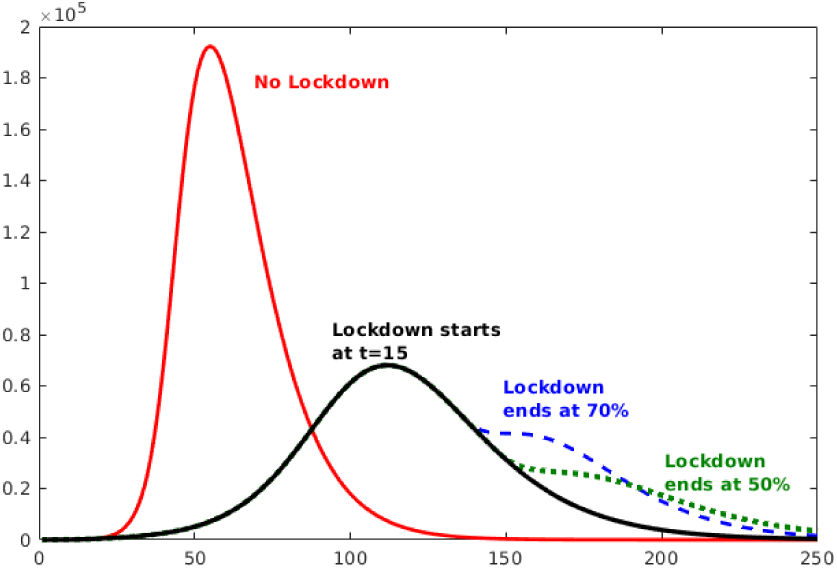
Comparison of the scenarios where the lockdown is relaxed after the percentage of people with visible symptoms (*O*) is reached the 70% and the 50% of the reported cases peak.

In general, we can observe several effects. The first effect is related to the timeliness of the lockdown, i.e. to the choice of anticipating *t*_Lock_. As expected, early lockdown reduces the height of the peak without much moving it forward in time. Conversely, lifting the lockdown too soon can make epidemic start again and reach values even higher than the ones before the release. A very peculiar and counter-intuitive effect can be generated if the lockdown is anticipated: in fact, a too early lockdown has the effect of delaying the start of the epidemic without attenuating its severity (see Sec. 8.2). Hence, early lockdown buy us some time but do only postpone the problem.

Another effect is represented by the impact of extreme quarantine measures on the post-lockdown time range. Indeed, increasing the strength α of the lock-down (i.e., reducing the social contacts) not only delays the time at which the lockdown can be lifted, but also induces a stronger reprise of the epidemic in the post-lockdown (see Sec. 8.2). Moreover, such approach could can result not sustainable for an economic system and motivate a gradual lifting of the lockdown measures to lessen the extant of further peaks. Furthermore, another counter-intuitive effect must be considered. Since to an attenuation *α* corresponds an effective reproduction number 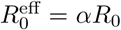, at the critical value *α*_crit_ = 1*/R*_0_ the epidemic neither grows nor decreases. Thus, after *t*_Lock_ the sys-tem stays stationary until the quarantine is released at *t*_Unlock_, and at this point the epidemic starts growing again as it was before the lockdown. In general, if *α < α*_crit_ the system looks to ameliorate (infected, hospitalized, all the infective compartments go down) but as soon as the lockdown is lifted, the epidemic starts again to reach its full extent (see Sec. 8.2). Nevertheless, our estimate *α ∼*0.5 *> α*_crit_ *∼*0.3 for the Italian lockdown gives us hope that, perhaps, it will not be necessary to follow a repeated seek-and-release strategy in the post-lockdown phase. On the other hand, if a lockdown strength *α ∼α*_*y*,_ can be attained without disrupting the economy, the epidemic could be contained until the creation, production and distribution of a vaccine.

## 5 Regional Scenarios

Starting with the first confirmed cases in Lombardy on 21 February, by the beginning of March the Covid-19 outbreak had spread to all regions of Italy. While the delay in the beginning of the infection is due to the different interaction among regions, once an epidemic has started it grows exponentially and the intake of external infected people becomes quickly irrelevant (see Sec. 8.6). As a consequence, the growth curves of the epidemic variables should tend to the same shape (see Sec. 8.5). Coherently, looking at the regional infographics released by the Italian National Healthcare Institute (ISS) [19], one may notice that they have a similar shape but different starting time (see Fig. 4). Such observation can be justified as follows: Italian regions are independent administrative units, where most of the population tend to work inside the resident region [29]. Hence, epidemics propagate from region to region via the fewer inter-regional exchanges (notice that Lombardy is among the Italian regions most involved in international trade connections [30], hence it appears as one of the most probable candidate for the start of the Italian epidemic). More practically, we estimate these delays by minimizing the distance among the observed curves (see also Sec. 8.4); results are reported in Tab. 1. Notice that, assuming that Lombardy has been the first region (i.e. delay=0), the resulting regional delays are mostly correlated to geographical distances.

**Tab. 1.** Regional delays (in days)

|  |  |  |  |
| --- | --- | --- | --- |
| Lombardia | 0.0 | Molise | 10.6 |
| Emilia Romagna | 3.1 | Umbria | 11.8 |
| Marche | 4.3 | Abruzzo | 13.1 |
| Veneto | 5.7 | Lazio | 14.5 |
| Valle d’Aosta | 6.4 | Campania | 15.0 |
| P.A. Trento | 6.6 | Puglia | 15.7 |
| P.A. Bolzano | 8.0 | Sardegna | 16.2 |
| Liguria | 8.1 | Sicilia | 16.6 |
| Friuli Venezia Giulia | 8.9 | Calabria | 17.2 |
| Piemonte | 9.0 | Basilicata | 19.2 |
| Toscana | 10.4 |  |  |

**Tab. 2.** POLYMOD matrix aggregated for three age classes: *Y* oung (00 *−*19), *M* iddle (20 *−*69) and *E*lderly (70+).

|  | Y | M | E |
| --- | --- | --- | --- |
| Y | 2.35 | 0.44 | 0.67 |
| M | 0.47 | 0.59 | 0.50 |
| E | 0.50 | 0.55 | 0.80 |

**Fig. 4.**
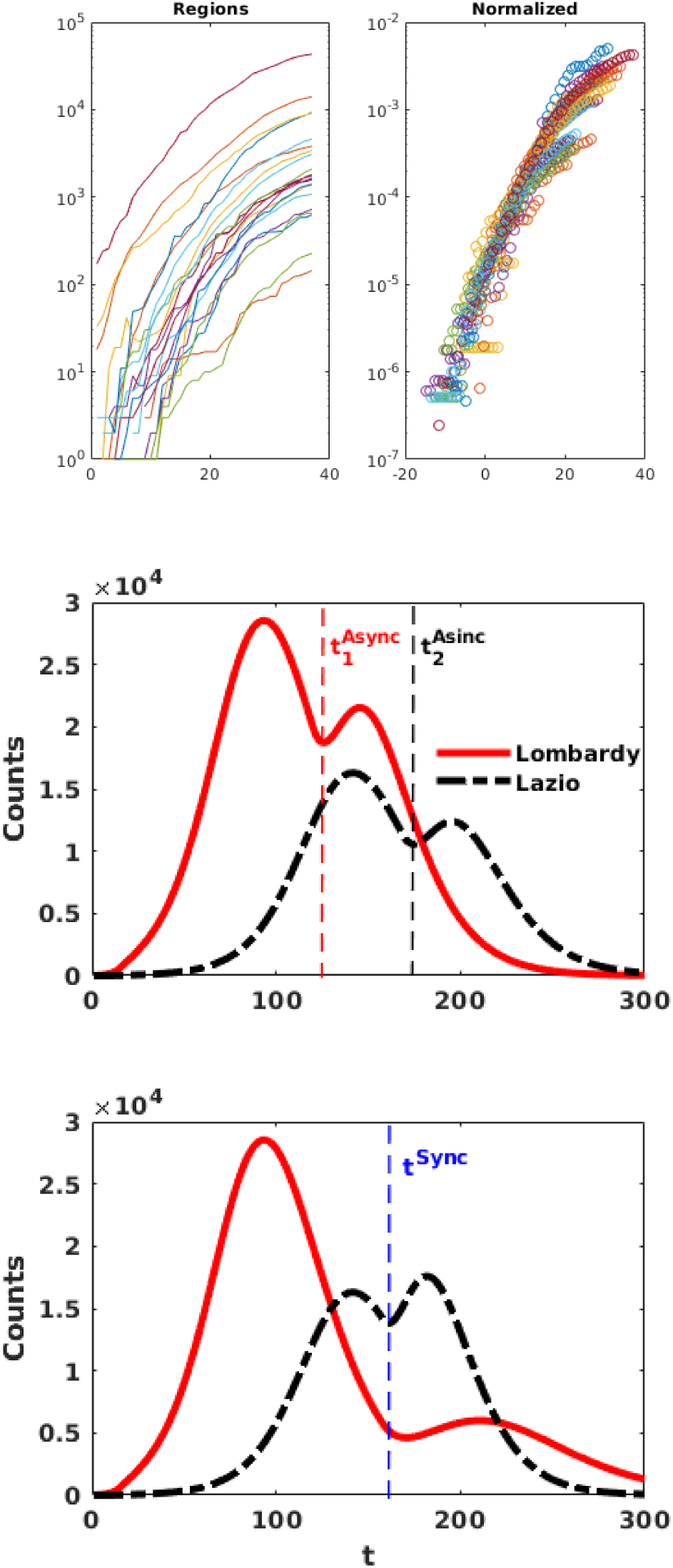
Upper panels: analysis of time delays among the start of epidemics in different regions (see Tab. 1). Lower panels: sketch of an Async(hronous) exit strategy (i.e. each region releases the lockdown following a given policy) respect to a Sync(hronous) exit strategy (i.e. the lockdown release follows the same policy, but applied to a nation wide scale). In particular, *t*^*Sync*^ corresponds to releasing the lockdown in all the region after the peak has fallen by 30%, while 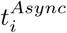 corresponds to releasing the lockdown in the *i*^*th*^ region after the peak *of such region* has fallen by 30%.

We assume that the Covid-19 outbreak spreads independently in each region of Italy; as argued before, such an approximation is reasonable after the epidemic has started and is even more accurate under lockdown conditions. Hence, we apply the parameters for the whole Italy to regional cases, where now the maximum number of individuals *N*_*i*_ is the population of the *i*^th^ region. Then, by summing up all the *S*_*i*_, …, *R*_*i*_, respectively, we obtain the evolution of Covid-19 epidemic throughout Italy. To evaluate the effect of heterogeneity in time delays, we compare the number of daily cases 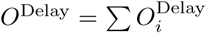 (obtained by taking into account the regional delays *t*_*i*_ as reported in Tab. 1) with the number of daily cases 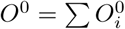 we would observe by considering the epidemics starts at the same time *t*_0_ in all regions. As expected, heterogeneity flattens the curve and shifts its maximum later in time. This is a first source of errors when fitting with a global model an heterogeneous dynamics.

Assuming that the right approach is the one with regional delays, we consider two possible exit strategies: in the first, that we call the *Async*hronous scenario, each region *i* lifts the lockdown at the time 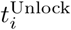 when the peak of 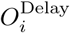 decreases by 30%; in the second, that we call the *Sync*hronous scenario, each region *i* lifts the lockdown at the same time *t*^Unlock^, i.e., when the global peak of *O*^Delay^ decreases by 30%. Notice that, since once started epidemics become essentially uncorrelated processes for each region, it could be safe to let each region have its own unlock time respect to having a global unlock. In Fig. 4 we compare the effect of a region-to-region release of the lockdown respect to releasing the lockdown at the same time in all the regions for the case of Lazio and Lombardy: in the latter case, there is a risk of having regions that have started earlier the contagion to wait a too much long time, and to have regions that have started later to have a too much strong rebound. Since not only epidemics, but also the ruin of an economy is a non-linear process, the *Sync* scenario can turn out to be even more disruptive than the epidemic itself (see also Fig. 3). Notice that analogous arguments hold - mutatis mutandis - also for the world/countries scenario.

## 6 Class Ages Scenarios

As we have already observed in the previous Section, heterogeneity strongly impacts on the results of a model [31]. Since the transmission coefficient is proportional to the contact rate of individuals, the rates of social mixing among different age classes could represent another important source of heterogeneity. This kind of information has been estimated either through large-scales surveys [24] or through virtual populations modeling [32]. While the POLYMOD [24] matrices have been extensively used to estimate the cost-effectiveness of vaccination for different age-classes during the 2009 H1N1 pandemic [33, 34], here we apply such matrices to the design of lockdown measures and exit strategies. Hence, to account for age classes, we extend our model by rewriting the transmission coefficient as *βC* (see Sec. 8.7), where *β* is the transmission probability characteristic of the disease, and *C* is the sociological matrix describing the contact patterns typical of a nation. For lack of further information, we assume *β* constant among age classes and *C* as in [24]. To simplify the analysis, we gather POLYMOD age groups into three classes: *Y* oung (00 *−*19), *M*iddle (20 *−*69) and *E*lderly (70+) (see Tab 2). Such aggregation puts together the most “contactful” classes (00 *−*19), the classes with the highest mortality risk (70+) [19], and the working class (20 *−*69).

Fig. 5 shows how the percentage of people with visible symptoms (*O*) varies once the age class heterogeneity has been added into the model. Introducing age structure in the model may lessen the peak we would observe in the case of full release and could allow to set up different exit strategies based on age-targeted policies for dampening a possible upturn of contagion. Specifically, quarantine measures applied to only elderly people may limit the impact of a renewed upward phase, while maintaining lockdown restrictions to all classes except the middle age class (20-69) could be enough to smooth and lessen the propagation of contagion in the post-lockdown phase.

**Fig. 5.**
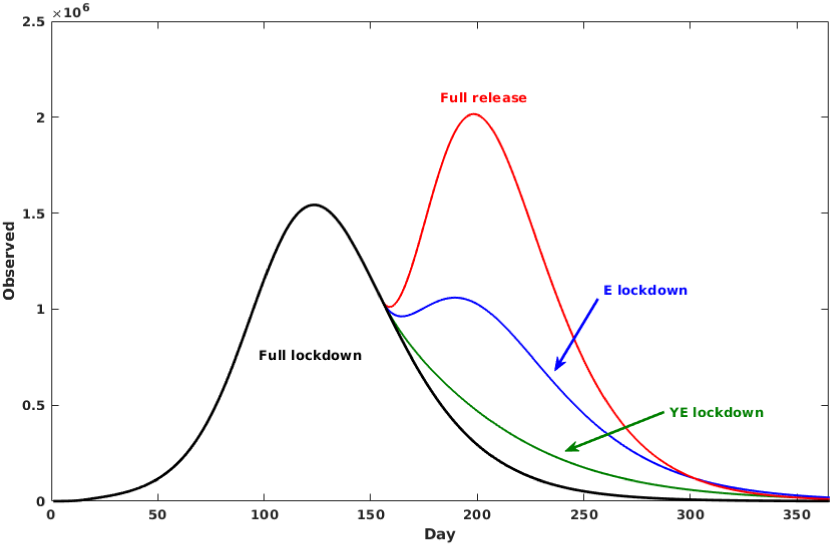
Comparison of the scenarios where the lockdown is relaxed only for particular age class with respect to a full release policy. Strategies: YE = quarantine young and elderly, E = quarantine elderly.

## 7 Conclusions

In this work we propose and test a generalized framework to study an epidemic contagion through a compartmental model based on geographical groups and age classes. We reveal how the promptness of lockdown measures has only a main effect on the timing of the contagion. More specifically, we show that strict social distancing policies reduce the extent of contagion during the lockdown period, but full recover occurs once such measures are relaxed. Interestingly, our study also contributes to uncover how local dynamics at regional level can be masked by observing the aggregate national system. Such regional heterogeneity, in fact, lowers and widens the curve of the contagion thus determining a shift forward in time for its peak at the aggregate national level. Moreover, by employing mobility data, we find that, due to the sparsity of interconnections across regions, contagion develops independently within each region once the epidemic is started. This, in turn, contributes to explain the delays observed in the alignment of the contagion curves across different geographical areas. Finally, we rely on the structure of social contacts to emphasize the role played by different age classes in the spreading of contagion. We notice that both young people (0-19) and elderly people (70+) are the most interconnected classes and therefore can have a greater influence on the post-lockdown phase. We show that while confirming quarantine policies to only elderly people can limit the possibility of a renewed upward phase in the contagion dynamics, relaxing the lockdown measures to only the middle age class (20-69) can be enough to smooth and lessen the propagation of contagion in the post-lockdown phase.

Although our study relies on data from relative to the Italian Covid-19 contagion, the proposed modeling approach can be broadly generalized to understand which dimensions, beside medical ones, should be considered to define leverage strategies for containing epidemics and mitigating its effects. Such a framework can also guide policymakers in the assessment of the trade-off between the impacts on the health care systems and the effects on wider economy. In particular, our analysis reveals how the timeline of post-lockdown measures can benefit from the inclusion of compartmental aspects, such as the geographical groups and age classes, in the decision process of a policymaker. For instance, this approach can be applied to evaluate specific quarantine measures for target age classes, such as those impacting on the most fragile age class (70+). For them, two competing policy paradigms could be applied: i) the first in which elderly people are grouped and confined to devoted residential buildings, thus increasing social mixing which enforces within connections and dismisses external relationships; ii) the second in which each elderly person is allocated alone thus limiting both intra and inter connections. Hence, in the first scenario in order to avoid the risk of spreading the contagion within a very fragile and concentrated community, strong social distancing measures with respect external individuals should be put in place. Such measures, if in the form of a strict lockdown applied regardless the age class membership, might have severe economic consequences and disruptive effects on production processes. Our approach thus sheds light on how target policy actions on specific age classes (namely, young and elderly people) can contain the spreading of contagion and keep a sufficient level of workforce at the same time.

## Data Availability

all the data used are freely available from online repositories or downloadable research papers

## Acknowledgements

A.S., E.B., M.C. and W.Q. acknowledge the support from CNR P0000326 project AMOFI (Analysis and Models OF social medIa) and CNR-PNR National Project DFM.AD004.027 “Crisis-Lab”.

## 8 Supplementary Information

### 8.1 Initial parameters estimation

In the early phases of the epidemic, observed quantities follow an approximately exponential growth *Y* ^Obs^ *∼Y*_0_*e*^*gt*^ as expected in most epidemic models. To understand what happens in our model, we notice that for *I/S «* 1 we can linearize Eq. 1 resulting in *I ∼I*_0_*e*^(*β−γ*)*t*^ and in *O ∼ργI*. Hence, minimizing the difference between *O* and *Y* ^Obs^ in such time range would yield estimates for *β, γ* such that *β −γ ∼g* and the basic reproduction number *R*_0_ *∼*1 + *g/γ* would increase linearly with the characteristic time *τ*_*I*_ = *γ*^*−*1^ for exiting the infective phase. Notice that most of the compartmental models based on ordinary differential equation will show an initial exponential growth phase with the same exponent (see Fig. 6); hence, in the early stage of the epidemic it is possible to successfully fit the “wrong” variables.

**Fig. 6.**
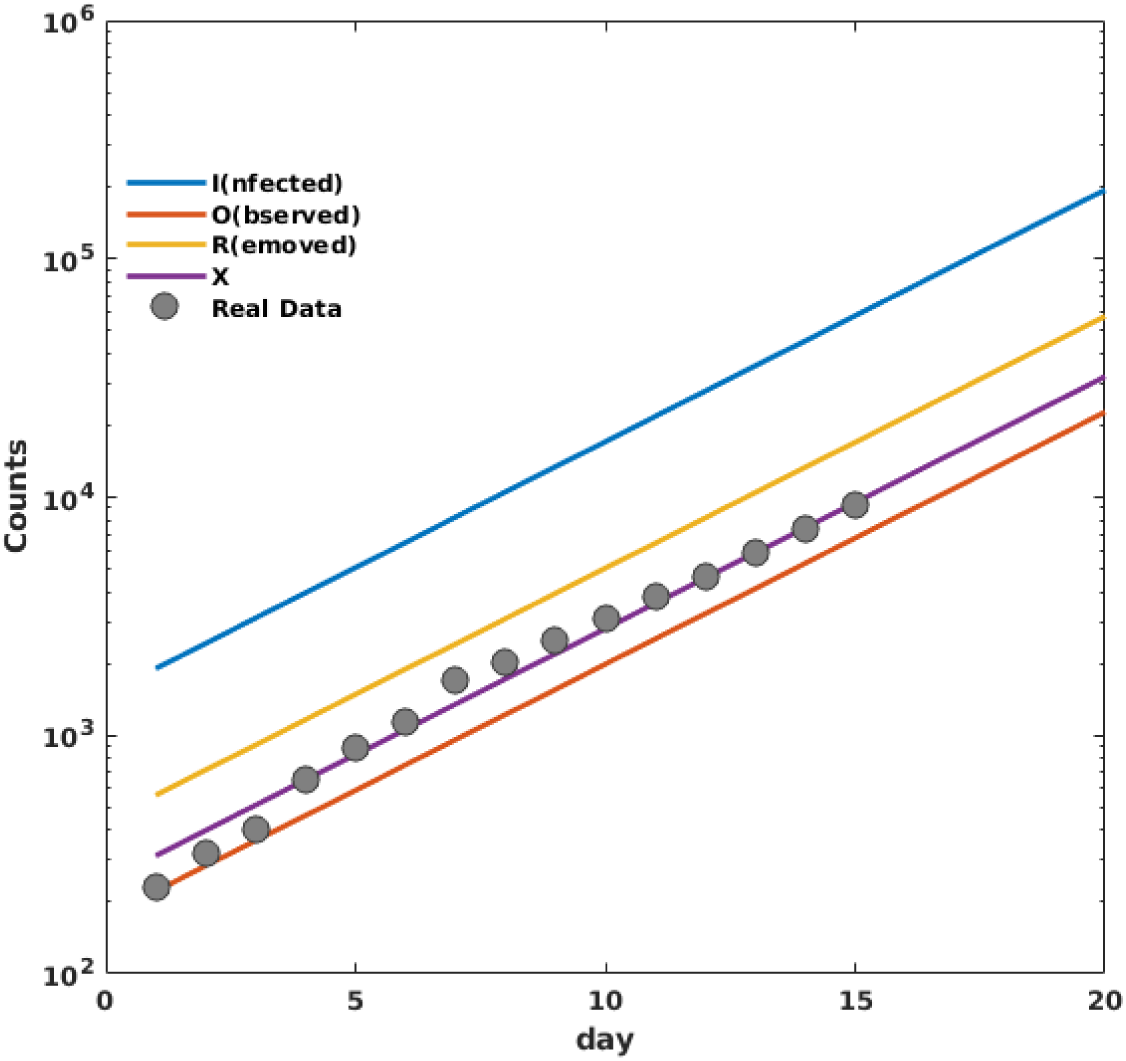
In the initial stage, most of the quantities experience an exponential growth with the same exponent; hence, it would be possibly to “successfully” fit the wrong variables. In the panel, we show the pre-lockdown growth of the number of *I*(nfected), *O*(bserved), *R*(emoved) individuals in our model (1). Full circles are the experimental counts of confirmed Covid-19 cases in Italy; *X* is the cumulative variable we use to fit the experimental data.

### 8.2 Effects of lockdown time and lockdown strength

By increasing the strength *α* of the lockdown (where *α* is the ratio between the trasmission *β* after and before the lockdown) the height of the peak lowers but shifts to farther times. On the other hand, slowing down the epidemic implies that lifting the lockdown would bring back the infection. In the left panel of Fig. 7, we show what happens by releasing the lockdown when the peak is fallen by 30%: stronger lockdowns induce a stronger reprise of the epidemic. An analogous effect can be observed by varying the lockdown time: anticipating the lockdown ameliorates the peak by decreasing its height, but shifts it to later time and retards the end of the epidemic.

**Fig. 7.**
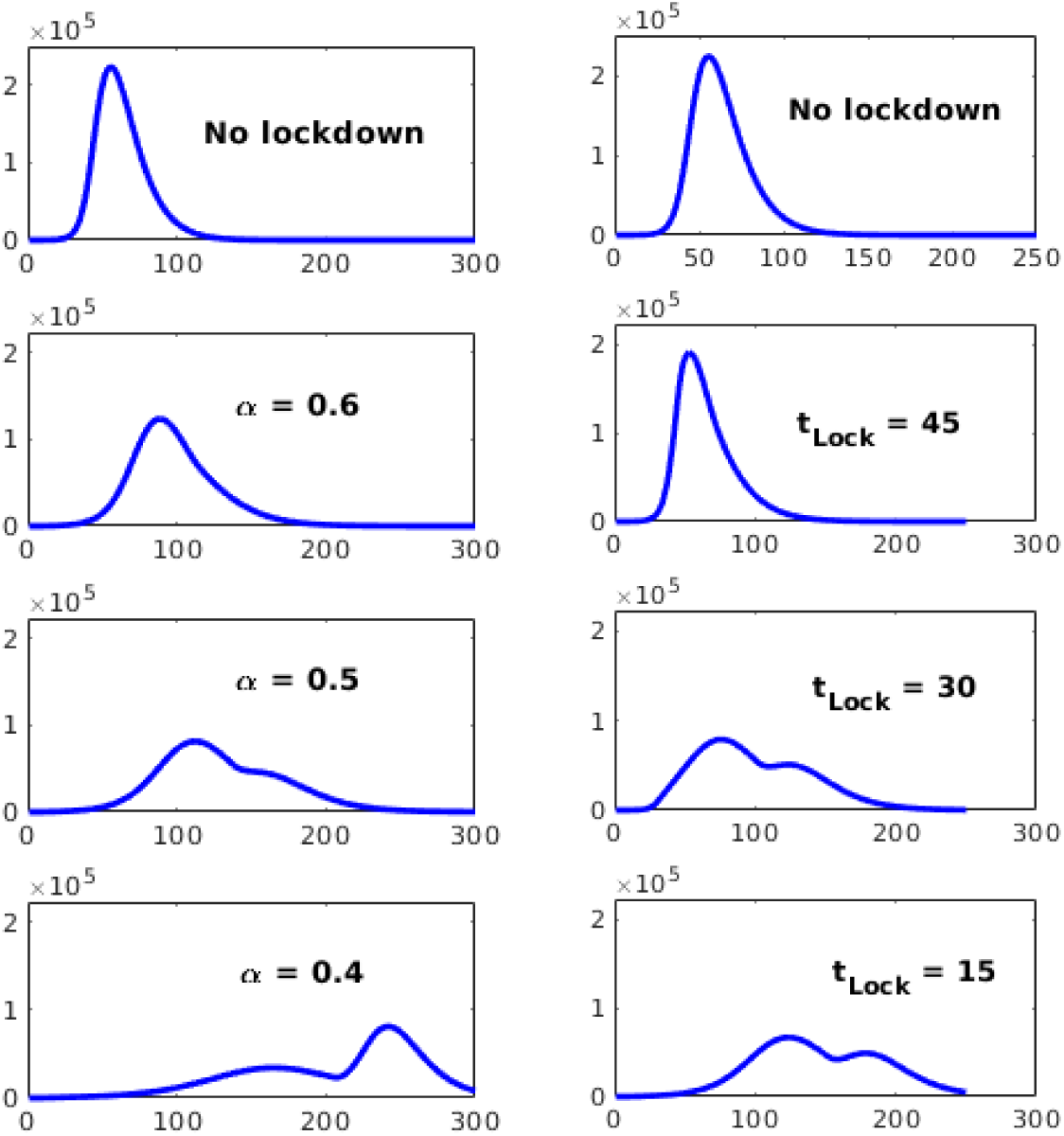
Left panel: variation of the behavior of the model by varying the lockdown strength *α*. Lockdown starts at *t*_Lock_ = 15 and is fully lifted when the peak has fallen by 30%. Right panel: variation of the behavior of the model by delaying the lockdown time *t*_Lock_. Lockdown strength is fixed at *α* = 0.5 and is fully lifted when the peak has fallen by 30%.

Contrary to what could be naively expected, an early imposition of the lockdown does not ameliorate the epidemics: in fact, anticipating too much the lockdown just shifts the timing of the epidemics, leaving its evolution unchanged (see Fig. 8). This is to be expected every time extreme measures of social distancing are applied in the very early, exponentially growing, stages. In fact, let us consider two countries *A* and *B*, having the same number of inhabitants, the same contact matrix, and the same number of infected people. If *A* and *B* decide to put a lockdown of strength *α* at time *t*_*A*_ and *t*_*B*_, respectively, at time *t* any quantity *y* would have grown as 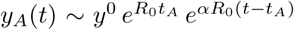 and as 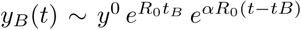 If there exists a *t* ′ such that *y*_*A*_(*t*) = *y*_*B*_(*t′*), the epidemics in *A* and in *B* will proceed in parallel (even in the non-linear phase) with a delay *t′ −t*. Therefore, if the epidemic dynamics of *A* and *B* are still well approximated by exponentials at times *<* max *{t, t′}*, then *t′ −t ∝* (*t*_*A*_ *−t*_*B*_), i.e, the country that has started the lockdown before will experience the same epidemic of the other country, just delayed in time. In particular, for identical initial conditions, we have that:

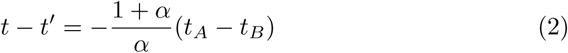

as long as all the times are before the initial exponential regime ends.

**Fig. 8.**
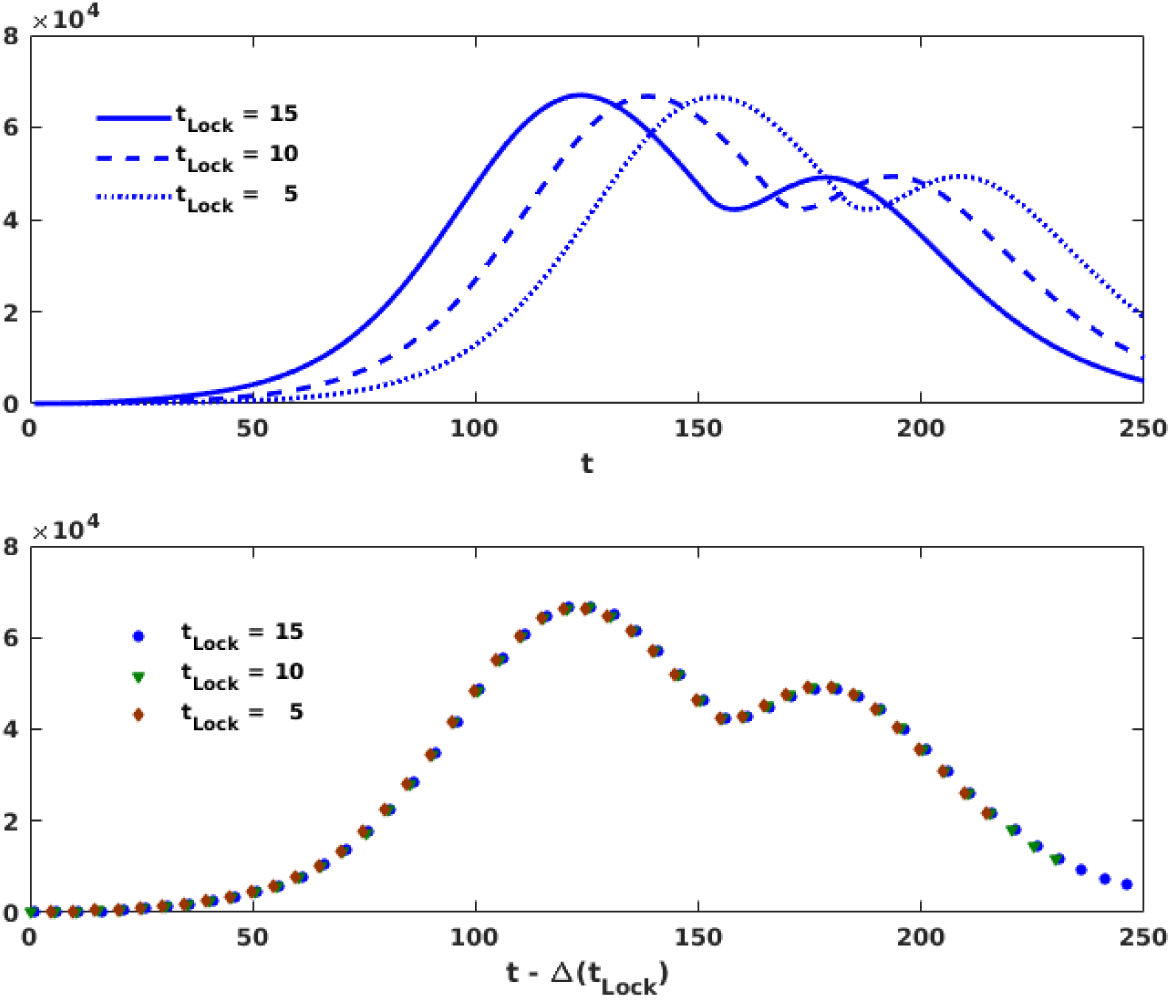
Upper panel: variation of the behavior of the model by anticipating the lockdown time. Notice that anticipating the lockdown leaves unchanged the behaviour of the epidemics, just shifting all the times of an amount proportional to how much the lockdown is anticipated. Lockdown strength is fixed at *α* = 0.5 and is fully lifted when the peak has fallen by 30%. Lower panel: by applying the Eq. 2, we show how the curves in the upper panel collapse on each other.

Finally, we notice that to each lockdown strength *α* corresponds an effective reproductive number 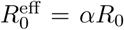 ; hence, for *α ∼α*_crit_ = 1*/R*_0_, the epidemics is expected to stay in a quiescent state where it does not either grow or decay sensibly. On the other hand, for *α < α*_crit_ the epidemics decreases; nevertheless, since this happens before a sufficient number of recovered individuals has built up herd-immunization, the height of the peaks after the lockdown lifting are almost unchanged if compared with the no lockdown scenario. Again, a “too good” intervention risks to postpone the problem without attenuating it. Notice that, if one applies lockdowns with *α < α*_crit_, it could be necessary to switch back and forth to lockdown to avoid the peak go beyond the capacity of a national healthcare system (see Fig. 9).

**Fig. 9.**
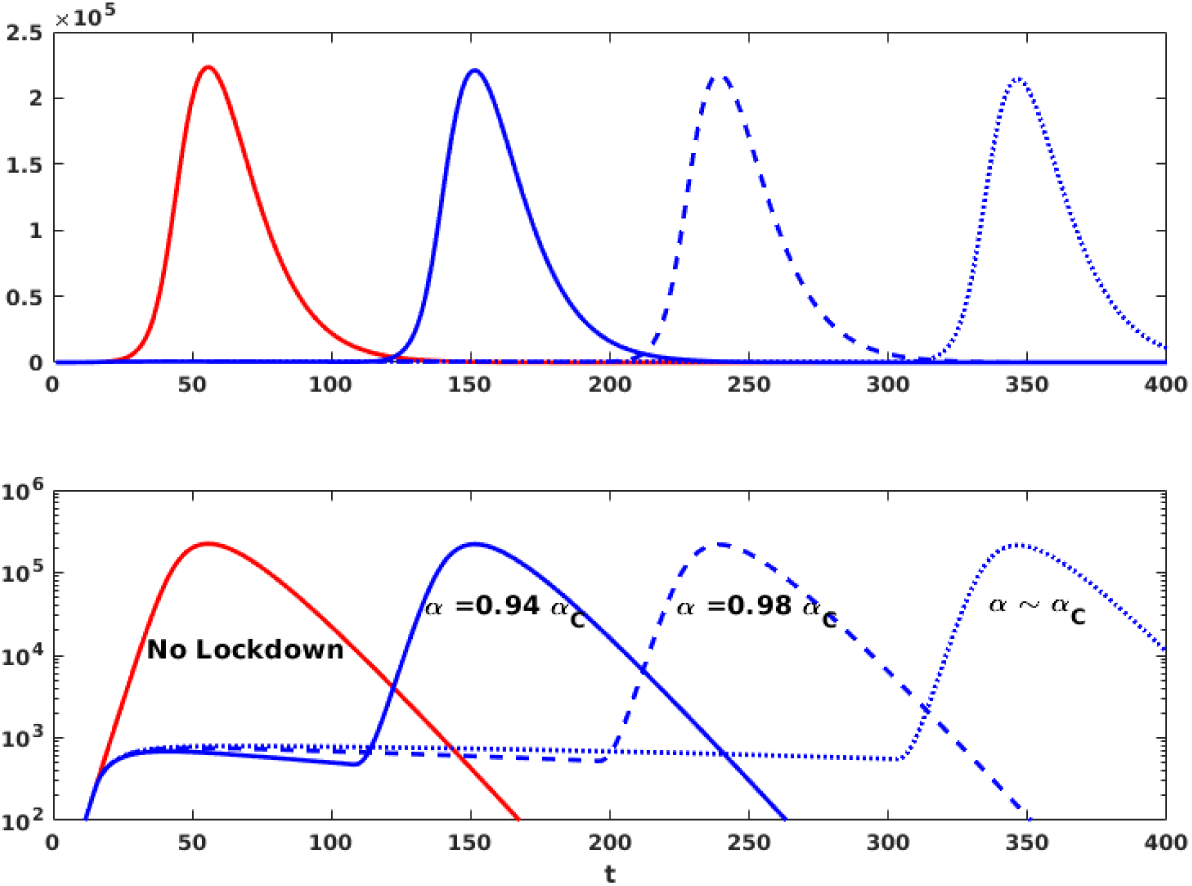
Upper panel: variation of the behavior of the model for lockdown strengths *α < α*_crit_ = 1*/R*_0_. Notice that the height of the peaks after the lockdown is released is almost unchanged if compared with the no lockdown scenario. Lockdown time is fixed at *t*_Lock_ = 15 and is fully lifted when the peak has fallen by 30%. Lower panel: for better clarity, the plot is also reported in log-linear scale.

### 8.3 Stationary state of the *SIOR* model

Let *X* = *O* + *R*. Then, *∂*_*X*_*S* = *−R*_0_*S* and 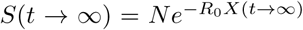. Since *O*(*t → ∞*) = *I*(*t → ∞*) = 0 and hence *R*(*t → ∞*) = *N −S*(*t → ∞*), we recover the same solution of the *SIR* model: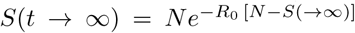. For *R*_0_ *∈* [2.5, 4.5], we have that the final fraction of uninfected population varies between 10% and 1%.

### 8.4 Estimation of the experimental time delays

We first normalize the observed data by dividing the number of non-zero observations in a region for the population of the region. Let *y*_*i*_ be the normalized observations for the *i*^th^ region. For each pair of regions *i, j*, we define the variation interval Δ_*ij*_ = [min_*ij*_, max_*ij*_] that contains the maximum number of points of both *y*_*i*_ and *y*_*j*_, i.e. min_*ij*_ = max*{*min(*y*_*i*_), min(*y*_*j*_)*}* and max_*ij*_ = min*{*max(*y*_*i*_), max(*y*_*j*_)*}*. The delay *t*_*ij*_ between the epidemics start in *i* and *j*, respectively, is calculated by minimizing the square norm of ‖ (Δ_*ij*_ *∩ y*_*i*_(*t*)) *\* (Δ_*ij*_ *∩ y*_*j*_(*t −t*_*ij*_)*‖*,where Δ_*ij*_ *∩ y* denotes the values of *y* falling in the interval Δ_*ij*_. Denoting with *T*_*i*_ the times corresponding to the observation in Δ_*ij*_ *y*_*i*_, it is easy to verify that *t*_*ij*_ = ⟨*T*_*i*_ ⟩ ⟨*T*_*j*_ ⟩, where ⟨ *T* ⟩ is the average value of the times contained in *T*.

### 8.5 Equivalence of normalized curves

Eq. 1 referred to region *k* becomes:

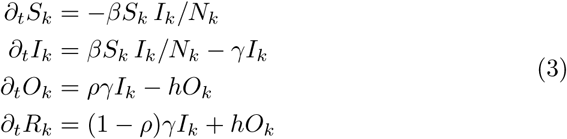

where *N*_*k*_ is the population of the region. By rewriting Eq. 3 in terms of normalized quantities *s*_*k*_ = *S*_*k*_*/N*_*k*_, … *s*_*k*_ = *S*_*k*_*/N*_*k*_, we obtain the same equation for all the regions:

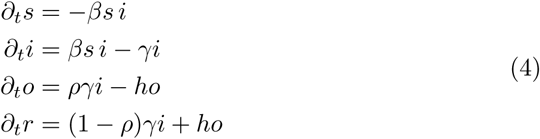

Hence, for similar initial conditions, by normalizing the experimental observations by the population, one should obtain similar time behaviors.

### 8.6 Regional metapopulation model

Let us assume that we know the fraction *T*_*kl*_ of people commuting from region *k* to region *l*, Eq. 4 becomes:

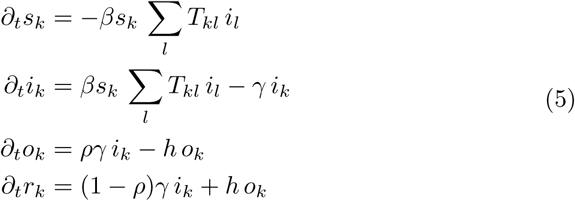

From mobility data, we know that ϵ_*k*_= ∑_*l≠k*_ *T*_*kl*_*/T*_*kk*_ *«* 1 and *T*_*kk*_ *∼*1; in particular, from Facebook mobility data we can estimate *⟨#x03F5;*_*k*_*⟩ ∼*10^*−*3^. If all the neighbors of a given region *k* are fully infected (i.e. *i*_*l*_ *=* 1 *∀ l / ≠ k*) and *i*_*k*_(*t*_0_) = 0, then the variation of *i*_*k*_ can be approximated as *∂*_*t*_*i*_*k*_ *∼ϵ*_*k*_ +(*β −γ*) *i*_*k*_. Namely, as soon as *i*_*k*_ *> ϵ*_*k*_, *i*_*k*_ will grow exponentially according to *∂*_*t*_*i*_*k*_ (*β γ*) *i*_*k*_ and ϵ_*k*_ will become irrelevant; that is to say, the dynamics of the regions will decouple. On the other hand, if epidemic is decaying everywhere, then *i*_*l*_ *«* 1 *∀ ≠ l k*; thus Σ_*l ≠k*_ *T*_*kl*_ *i*_*l*_ *« ϵ*_*k*_ and equation again decouple, having each region followed Eq. 4 separately. An alternative source of network information comes from the recorded movements of individuals. Both airline transportation network [35, 36] and individual work commutes [37, 38] have played important roles in understanding the spread of infectious diseases.

### 8.7 Social mixing

To take account for social mixing, we rewrite the transmission coefficient as the product of a transmission probability *β* times a contact matrix *C* whose element *C*_*ab*_ measure the average number of (physical) daily contacts among an individual in class age *a* and an individual in class age *b*. Notice that the probability that a susceptible in class *a* has a contact with an infected in class *b* is the product of the contact rate *C*_*ab*_ times the probability *I*_*b*_*/N*_*B*_ that individual in class *b* is infected. Hence, denoting with *S*^*a*^, …, *R*^*a*^ the number of *S*(usceptibles),…,*R*(emoved) individuals in class age *a*, we can rewrite Eq. 1 as:

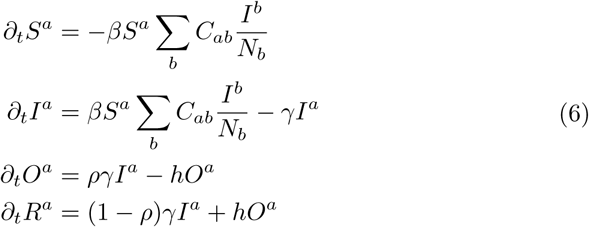

Although the form of Eq. 6 is similar to Eq. 3, here it is not possible to consider separate evolutions for the different age classes since, differently than the inter-regional mobility matrix *T*, the off diagonal elements of the social matrix *C*_*a,b*_, *a ≠ b*, measure the interaction among different age classes and are of the same magnitude of the diagonal elements *C*_*aa*_ measuring the interaction among individuals of the same age class.

Notice that Eq. 6 can be summed up, and the resulting equation can be obtained by substituting *β → βC*^eff^ in Eq. 1, where 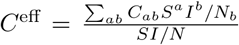 is the average contact value among infected and susceptible individuals of all age classes.

## Footnotes

1 See, e.g.: https://www.sciencemag.org/news/2020/03/mathematics-life-and-death-how-disease-models-shape-national-shutdowns-and-other

2 Those data are part of the Facebook project “Data for Good”, and illustrates where users, who allowed the social network to track their location, moved. See https://dataforgood.fb.com/docs/Covid-19/.

